# The effects of RYSEN-induced supportive, propelling and impeding forces on step parameters and muscular control in walking after stroke

**DOI:** 10.1101/2025.06.05.25329024

**Authors:** Sanne Ettema, Tom J.W. Buurke, Sina David, Coen A.M. van Bennekom, Han Houdijk

## Abstract

**Background:** Body weight support (BWS) devices are increasingly used for gait training after stroke. The RYSEN not only allows for vertical BWS but also for the addition of mediolateral and anteroposterior forces. To assess to what extent the RYSEN could be used for targeted gait training, this study aimed to investigate the effect of BWS with and without the addition of mediolateral and anteroposterior forces on step parameters and muscular control in individuals after stroke and able-bodied individuals.

**Methods:** In this cross-sectional study, fifteen individuals after stroke and fifteen able-bodied individuals completed several overground walking conditions: unsupported and BWS with(out) mediolateral and anteroposterior forces. Step length, step width (variability), single stance time and walking speed were determined using motion capture data. Muscular control was assessed using electromyography. Linear mixed-effect models were used to analyze the (interaction) effects of condition and group on step parameters. Statistical parametric mapping was used to analyze muscular control.

**Findings:** Providing BWS led to increased speed and balance confidence in individuals after stroke, and caused decreased speed, step width variability and m. Gluteus Medius activity in able-bodied individuals. Adding mediolateral forces did not lead to additional changes. Adding anterior forces increased speed in individuals after stroke. Adding posterior forces slightly increased plantar flexor activity during push-off in able-bodied individuals.

**Interpretation:** RYSEN BWS facilitates gait training at increased speed and balance confidence. Adding mediolateral or anteroposterior forces did not provide strong additional effects. Further research may determine how these forces can be better utilized during training.

## Introduction

Body weight support (BWS) devices are increasingly being used to improve walking ability in individuals after stroke (1, 2). Recently, the RYSEN (Motek Medical, Houten, the Netherlands) has been launched as an innovative multidirectional BWS device that enables overground walking in a 3D space (3). The device not only allows for position-independent vertical forces that provide BWS, but also for the addition of mediolateral and anteroposterior forces, which can be set independently of BWS through decoupled degrees of freedom (3). In line with the assist-as-needed principle, the direction and magnitude of the forces can be adjusted to an individual’s needs. To purposefully exploit the potential of the RYSEN for gait training, it is crucial to understand how different directions and levels of BWS and mediolateral or anteroposterior forces affect gait characteristics, for instance in individuals after stroke, who are considered a main target group of BWS gait training (1).

The effect of BWS on gait characteristics has been extensively investigated (4-7). BWS is known to mainly affect muscle activity and to a lesser extent spatiotemporal gait parameters, while kinematics are preserved (6). It has been argued that low levels of BWS (i.e., up to 30% unloading) enable individuals to maintain their natural gait (8, 9). Among numerous studies evaluating the effect of BWS on gait characteristics, the results show inconsistency (9). This is partly because various devices have been used, while the effects of BWS are considered device-specific (10). It has, therefore, been argued that optimal training settings have to be customized to each device separately. However, these have not yet been identified for the RYSEN.

The RYSEN offers the possibility of adding independent mediolateral forces on top of the vertically directed BWS forces. Using mediolateral forces, the RYSEN reacts to an individual’s lateral sway to restrict sideward movement with a contrary-directed force. As such, mediolateral forces may act as a lateral stabilizer by restricting lateral translations and enhancing frontal plane stability (3, 11). It is known that, whereas sagittal plane stability can be largely maintained through passive dynamics of the limbs, frontal plane stability requires active neuromuscular control (12, 13). Experimental studies have shown that lateral stabilizers without BWS enhance gait stability (11, 14) and reduce the metabolic cost of walking (15), for instance, due to reduced needs for muscular control to fine-tune balance after foot placement (16) and reduced external mechanical work because of decreased step width (17). Similarly, mediolateral forces of the RYSEN could potentially reduce the load of walking in addition to BWS and selectively be used to set the level of challenge for balance control.

Additionally, the RYSEN allows for the application of anteroposterior forces on top of BWS. These forces can act in or against the walking direction and either propel (i.e. anterior forces) or impede (i.e. posterior forces) gait. Previous studies on anteroposterior forces without BWS have shown that anterior forces increase walking speed and therefore allow walking with reduced propulsion capacity, but increased braking impulses (18, 19). On the other hand, posterior forces are known to decrease braking impulses and challenge the user to increase propulsion capacity (18, 20, 21). However, the effects of anteroposterior forces on top of BWS have not been studied extensively. In a recent study, anteroposterior forces in the RYSEN were found to affect walking speed and gait kinetics in healthy individuals (10). Still, the effects on gait in clinical target populations of the device, e.g. individuals after stroke, are not yet understood.

The RYSEN has been invented for clinical applications to aid gait training in, amongst others, individuals after stroke and could potentially improve gait stability (22), walking speed (23) and propulsion capacity (24). To assess to what extent the RYSEN could be used for targeted gait training, this study investigated the effect of BWS with and without the addition of mediolateral, and anteroposterior forces on step parameters and muscular control in individuals after stroke and able-bodied individuals. We hypothesized that BWS and the addition of mediolateral forces would enhance gait stability, reflected by reductions in step width, step width variability (25) and the activity of muscles used to control balance (15, 16). Furthermore, we hypothesized that anterior forces would induce walking at increased speeds. Additionally, we expected that posterior forces would cause individuals to increase the activity of muscles that are associated with propulsion capacity to maintain walking speed.

## Methods

### Participants

Seventeen individuals after stroke and fifteen able-bodied individuals provided written informed consent to participate in this study. Inclusion criteria were age > 35 years, weight < 135 kg, length < 2 m, the ability to walk at least 10 m without assistance or walking aids, and the ability to communicate with the experimenters. Individuals after stroke were allowed to use their ankle-foot orthosis during the experiment if needed and to use a walking aid for manual support (26) but instructed not to use it for weight-bearing assistance.

Able-bodied individuals were recruited through personal communication and individuals after stroke by convenience sampling in Heliomare rehabilitation center between 23 October 2023 and 28 June 2024. Sample size calculation was based on a comparable study on BWS (27) that found a ƞ^2^ of 0.16. Combined with an α error probability of 0.05 and a β power of 0.95, a sample size of 13 individuals per group was required (computed by G*Power, version 3.1.9.7) (28). Because of the high heterogeneity of individuals after stroke, this number was increased to 15 individuals per group.

The procedures of this study were consistent with the Declaration of Helsinki (29) and approved by the Medical Ethics Review Board of the University Medical Center Groningen (2022/220). This study reporting follows the STROBE guidelines for reporting cross-sectional studies (Appendix A).

### Materials

The RYSEN BWS system (3) (Motek Medical, Houten, The Netherlands) unloaded participants during overground walking. Participants wore a harness that was strapped to the body at the abdomen, pelvis and upper legs. The harness was attached to an overhead sling, which was connected to rails at the ceiling through four cables. Each of the cables was controlled via an actuator to move with the participant and to ensure that the resultant forces experienced by the participant stayed constant, independent of position in space. The setup allowed participants to walk in an area of 13.5 x 3.5 m.

To assess spatiotemporal gait parameters, position data from two markers attached to the heel of each foot and a marker attached to the sacrum were recorded with an Optotrak motion analysis system (Nothern Digital Inc, Ontario, Canada) at 100 Hz sampling frequency. Surface electromyography (EMG) data (FREEEMG, BTS Bioengineering, Milan, Italy) were recorded bilaterally at 1000 Hz sampling frequency on the following lower limb muscles: m. Gluteus Medius, m. Semitendinosus, m. Rectus Femoris, m. Vastus Lateralis, m. Gastrocnemius Medialis, m. Soleus and m. Peroneus Longus and m. Tibialis Anterior according to SENIAM guidelines (30).

### Procedures

The protocol consisted of overground walking up and down a 10 m space at preferred speed in four blocks: unsupported walking (UW), BWS, BWS+ML, and BWS+AP, all separated into multiple conditions (Figure 1). During UW, participants walked without the RYSEN. During the RYSEN conditions, BWS and anteroposterior forces were expressed in percentages of body weight. The magnitude of mediolateral forces was not adjustable, as they served solely to fix the sling bar in mediolateral direction. During BWS, participants walked with 10% and 30% BWS. During BWS+ML and BWS+AP, participants walked again with 10% and 30% BWS, combined with either mediolateral (+ML), anterior (+1%, +2% body weight) or posterior (- 1%, -2% body weight) forces. The trial duration was 1.5 minutes for able-bodied individuals and 1 minute for individuals after stroke to ensure they could finish the protocol. For each individual, the blocks (i.e. BWS, BWS+ML and BWS+AP) as well as the conditions within each block were offered in randomized order. For practical and safety reasons, individuals after stroke always started with UW and able-bodied individuals started or ended with UW. After each trial, individuals after stroke were asked to rate their balance confidence on a 0 to 100% rating scale (31). All individuals after stroke had received at least one training session with the RYSEN as part of their inpatient rehabilitation program before participation in this study.

**Figure 1.**
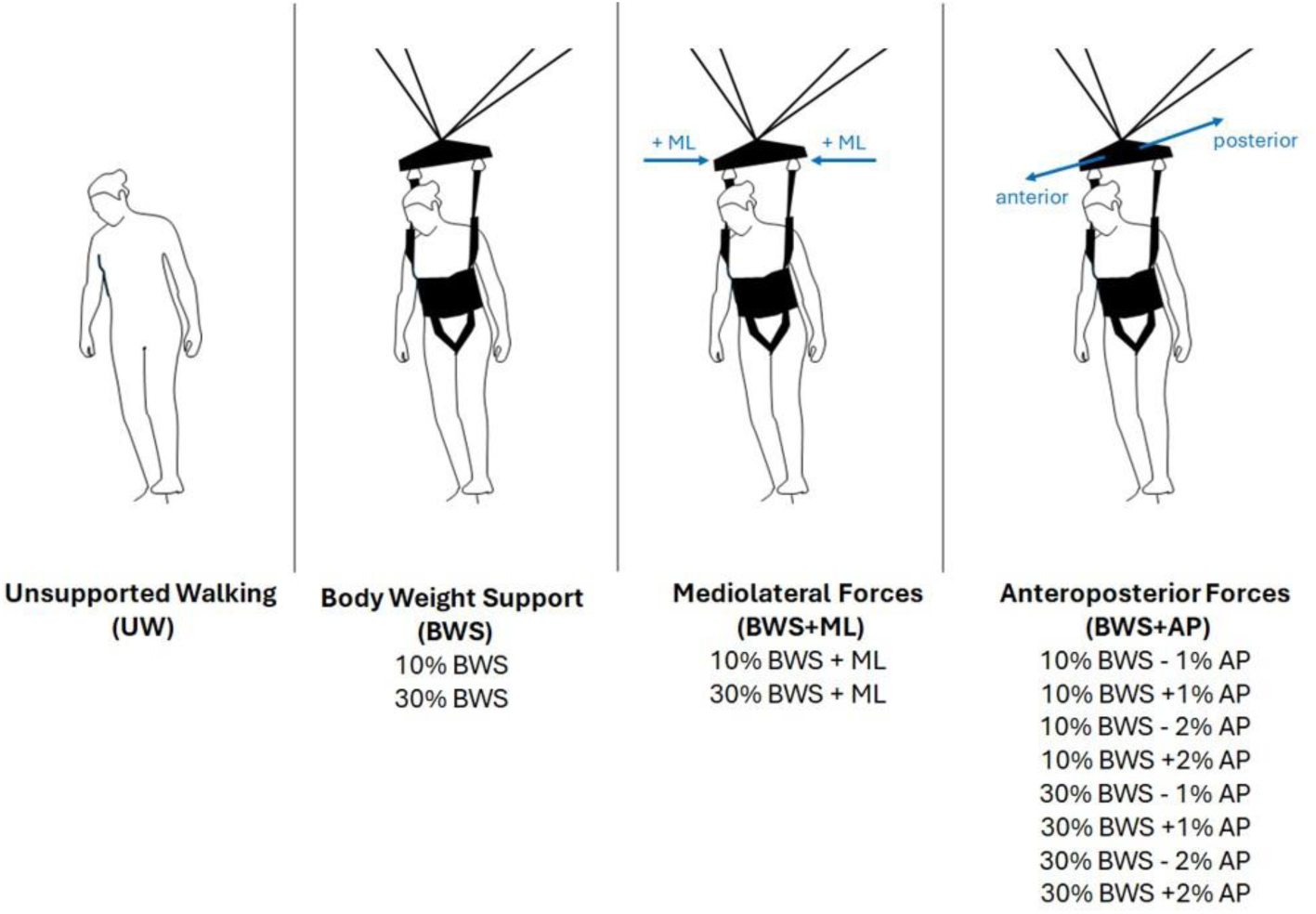
Experimental protocol consisting of four blocks (Unsupported Walking (UW), Body Weight Support (BWS), BWS + Mediolateral (ML) Forces, BWS + Anteroposterior (AP) Forces), separated into multiple conditions. The protocol was performed in block randomized order.

### Data analysis

Data were analyzed in MATLAB (version r2023a; The Mathworks Inc., Natick, MA, USA). Marker position data were used to detect gait events with a coordinate-based algorithm that finds local peaks and valleys in the anteroposterior direction of foot marker data, corresponding to heel-strikes and toe-offs (32).

Global marker position data of the sacrum were used to define a local frame, in which the x-axis was enforced to be oriented in the walking direction, defined as the direction of the sacrum velocity. Data were filtered beforehand using a moving average filter of 1 and 5 s for able-bodied individuals and individuals after stroke respectively to correct for step-to-step pelvis rotations that could affect the determination of the walking direction. Different time spans accounted for step time differences between groups. Step length and step width were expressed in the local frame. Step length (m) was calculated as the anteroposterior distance between both heel markers at the moment of heel strike. Step width (m) was calculated as the mediolateral distance between both heel markers at the moment of heel strike. Step width variability (m) was calculated as the standard deviation of step widths within each condition. Single stance time (s) was calculated as the difference in time between toe-off of the ipsilateral and heel-strike of the contralateral leg. Walking speed (m/s) was calculated by taking the time derivative of the position data of the sacrum. To correct for habituation, the first ten seconds were left out of the analysis for walking speed and the first three strides were excluded from the analyses for step length, step width and single stance time. Moreover, the first stride after a directional change was left out to ensure directional changes did not affect the determination of the walking direction.

EMG data were band-pass filtered (2^nd^ order Butterworth, bidirectional between 20 and 250 Hz). Thereafter, EMG signals were rectified and low-pass filtered (4^th^ order Butterworth, bidirectional at 10 Hz) to obtain envelopes. For each individual, EMG data of each muscle were normalized to the peak value over all conditions and time-normalized to gait cycles from heel-strike to heel-strike. Gait cycles of the left and right leg were combined for able-bodied individuals, as EMG is known not to differ substantially between both legs (33). For individuals after stroke, the most and least affected leg were analyzed separately.

### Statistics

To determine the effects of BWS, BWS+ML and BWS+AP on step parameters, differences compared to UW were assessed using linear mixed-effect models for each of the three blocks separately.

Two statistical models were used, depending on whether the assessment concerned a comparison between *legs* (i.e. most affected leg, least affected leg, able-bodied leg) or *groups* (i.e. stroke or able-bodied). In both models, the fixed effects were defined as age, condition and *leg* or *group,* and a random intercept was used for each participant.

The assessment of step length and single stance time concerned a comparison between *legs*, using the model below, with the most affected leg as a baseline:

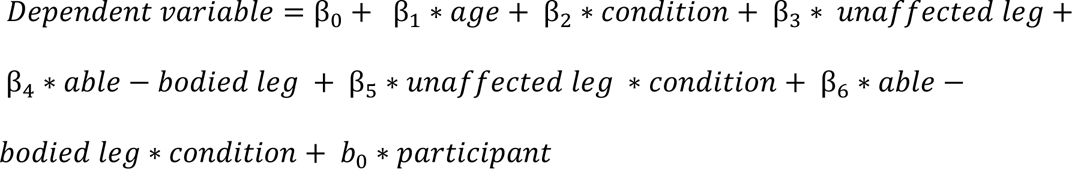

The assessment of step width (variability) and walking speed concerned a comparison between *groups*, using the model below, with individuals after stroke as a baseline:

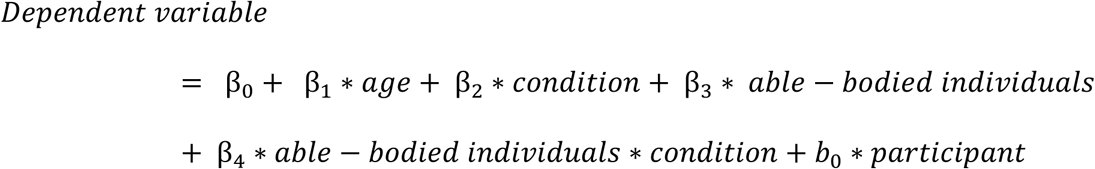

To determine the effects of BWS, BWS+ML and BWS+AP on muscular control, differences compared to UW were assessed using 1D Statistical parametric mapping (SPM) repeated measures ANOVA (34) for each block separately. Separate analyses were performed for the most affected leg of individuals after stroke and the able-bodied controls. Post-hoc testing was performed using SPM 1D dependent t-tests and only significant post-hoc effects overlapping with the main effects were interpreted to correct for multiple comparisons.

Differences in confidence in balance control were assessed using repeated measures ANOVA for each block separately and followed by dependent t-tests when significant. Statistical significance was set at *p* <0.05 for all tests.

## Results

Fifteen individuals after stroke and fifteen able-bodied individuals were included in the analyses (Table 1). Two individuals after stroke were excluded because they did not succeed in walking in the RYSEN without the experimenter’s assistance.

**Table 1.**
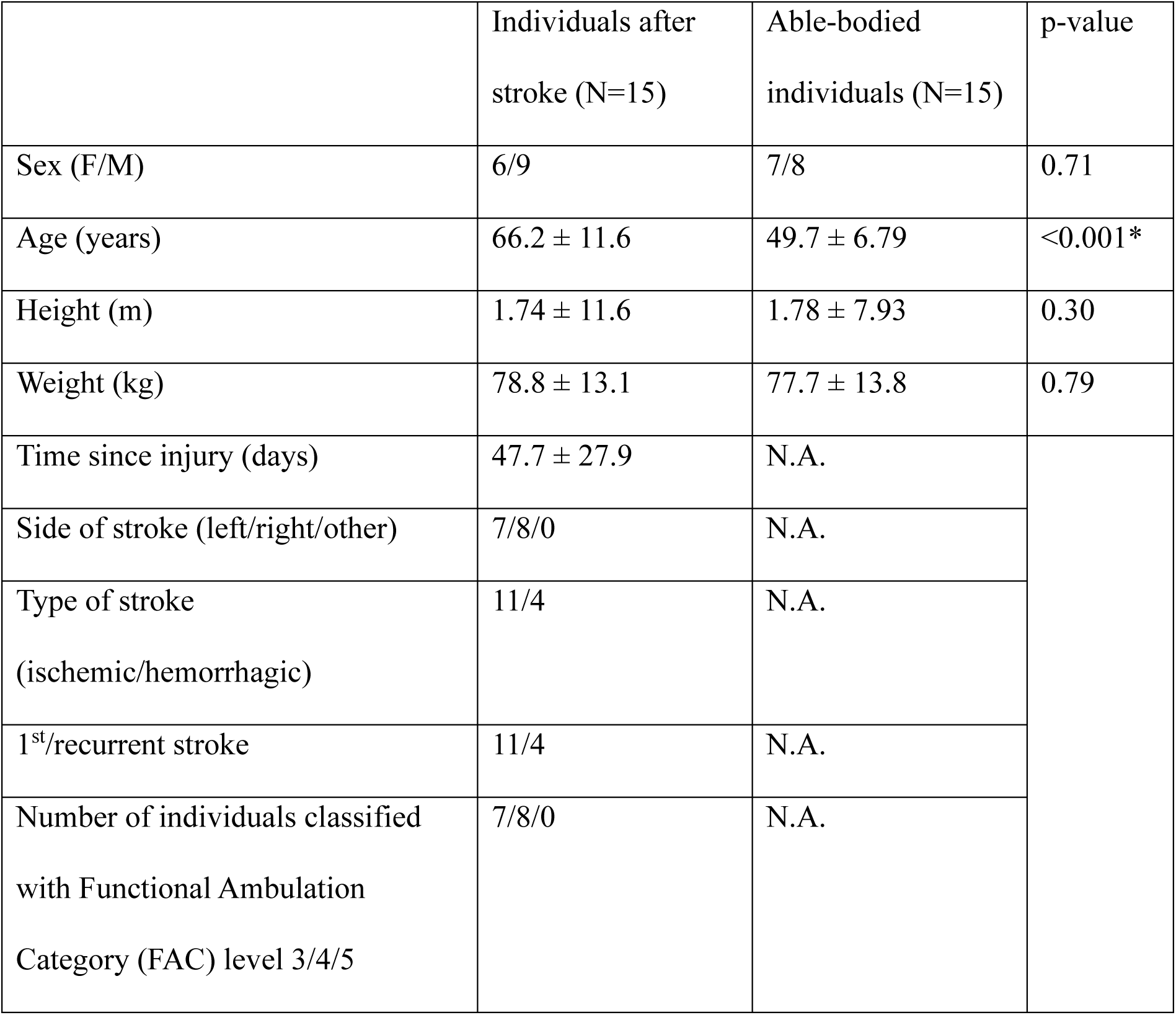
Characteristics of the study population.

The following sections present the results for each experimental block (i.e. BWS, BWS+ML and BWS+AP) respectively. First, step parameters will be discussed including the significant effects of *leg* or *group*, condition, and their interaction. Thereafter, significant post-hoc EMG results will be discussed.

For all three blocks, step length (p<0.001) and walking speed (p<0.001) were larger for able-bodied individuals compared to individuals after stroke. For BWS and BWS+ML, step width was larger for individuals after stroke than for able-bodied individuals (p<0.05). Detailed statistical outcomes are shown in Appendix B.

### Body weight support conditions

Walking speed increased with 10% BWS (p=0.01) and 30% BWS (p=0.03, Figure 2) compared to UW. A significant condition*group interaction with 10% BWS showed that walking speed increased for individuals after stroke and decreased for able-bodied individuals (p<0.001). Step length increased with 10% BWS (p=0.002) and 30% BWS (p=0.002). Single stance time increased with 30% BWS (p=0.01). For step width variability, significant condition*group interactions were found, indicating that step width variability increased in individuals after stroke and decreased in able-bodied individuals with 10% (p=0.003) and 30% BWS (p=0.02).

**Figure 2.**
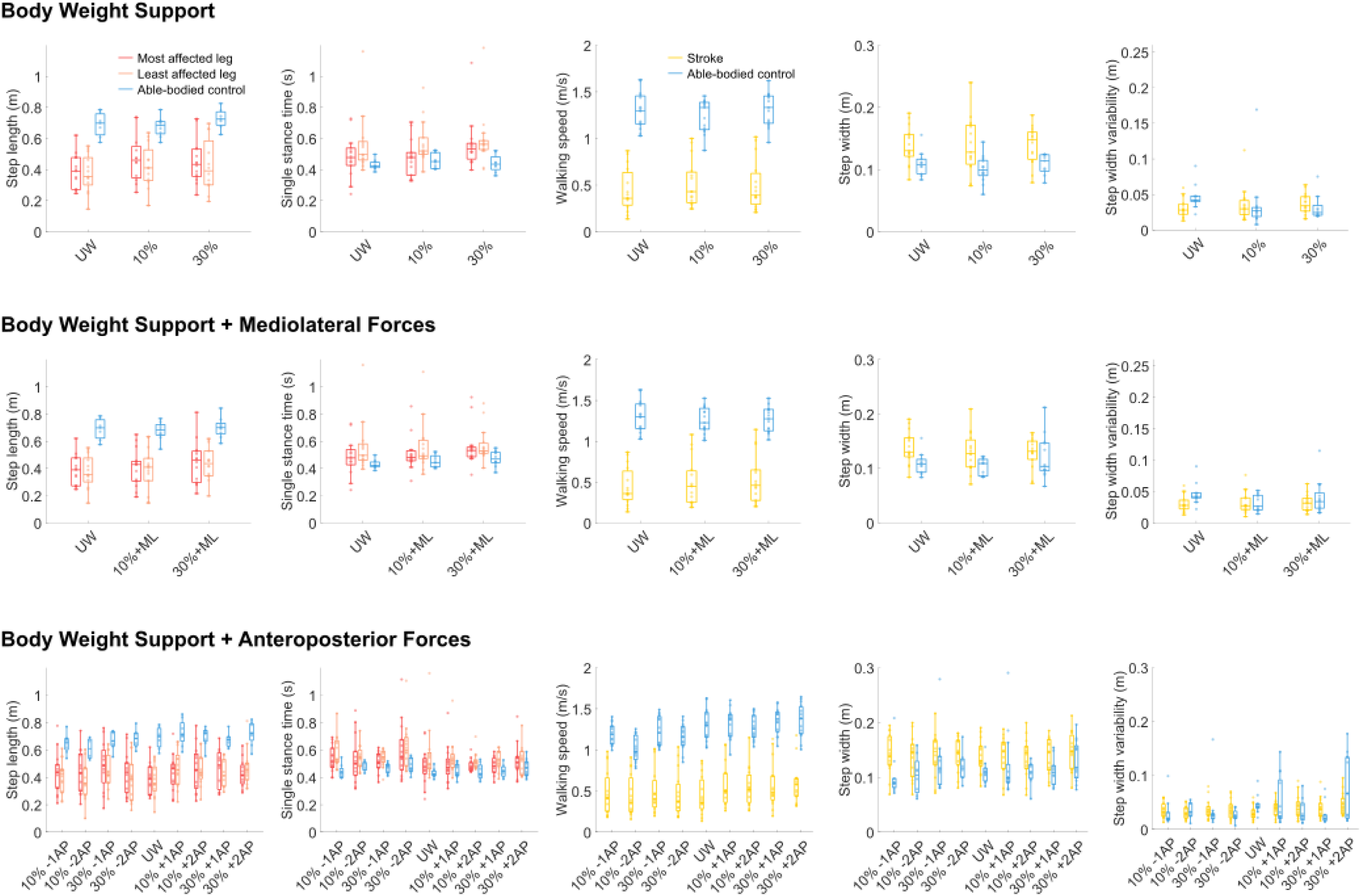
Step parameters for Unsupported Walking (UW), Body Weight Support (BWS, upper panel), Mediolateral Forces (ML, middle panel) and Anteroposterior Forces (AP, lower panel). The boxplots show the following: box, interquartile range (IQR, 25^th^ – 75^th^ centiles); upper whisker, upper adjacent; lower whisker, lower adjacent; line, median. Individual data points are presented as transparent dots.

For able-bodied individuals (Figure 3), decreased m. Gluteus Medius activity (p<0.001) was found with 30% BWS (p<0.001). For m. Tibialis Anterior, decreased activity was found during midstance with 10% BWS (p=0.01) and increased activity during mid-swing with 30% BWS (p=0.02, p=0.01).

**Figure 3.**
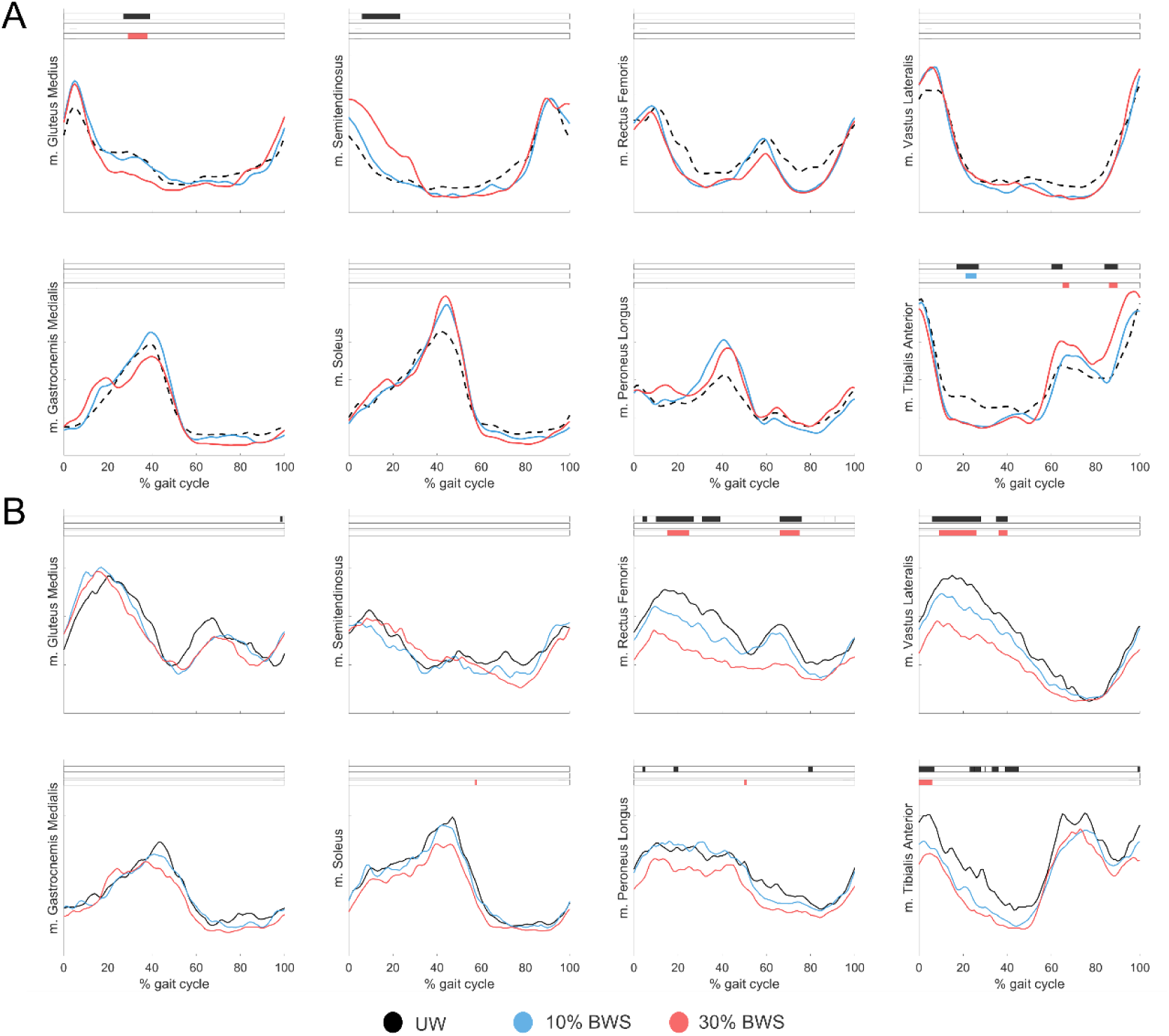
Mean muscle activation patterns across the gait cycle during walking with Body Weight Support (BWS) for *A)* able-bodied individuals and *B)* individuals after stroke during Unsupported Walking (UW; black), 10% BWS (blue) and 30% BWS (red). Black (main effect) and colored (post-hoc effect) boxes show the part of the gait cycle in which a significant difference is present (p<0.05).

For individuals after stroke (Figure 3), decreased m. Rectus Femoris activity was found during midstance and initial swing with 30% BWS (all p<0.001). For m. Vastus Lateralis, decreased activity was found during the stance phase with 30% BWS compared to UW (p<0.001, p=0.01). For m. Tibialis Anterior, decreased activity during loading response was found with 30% BWS compared to UW (p<0.001).

### Body weight support with mediolateral forces

Walking speed was higher with 30%+ML compared to UW (p=0.01, Figure 2). There were significant condition*group interactions, indicating that walking speed increased for individuals after stroke and decreased for able-bodied individuals with 10%+ML (p=0.02) and 30%+ML (p=0.01). Step length increased with 30%+ML compared to UW (p=0.02). Single stance time increased with 10%+ML (p=0.04) and 30%+ML (p=0.03). These results were comparable to those found when walking with BWS only.

For able-bodied individuals (Figure 4), decreased m. Tibialis Anterior activity was found with 30%+ML during stance (p<0.001).

**Figure 4.**
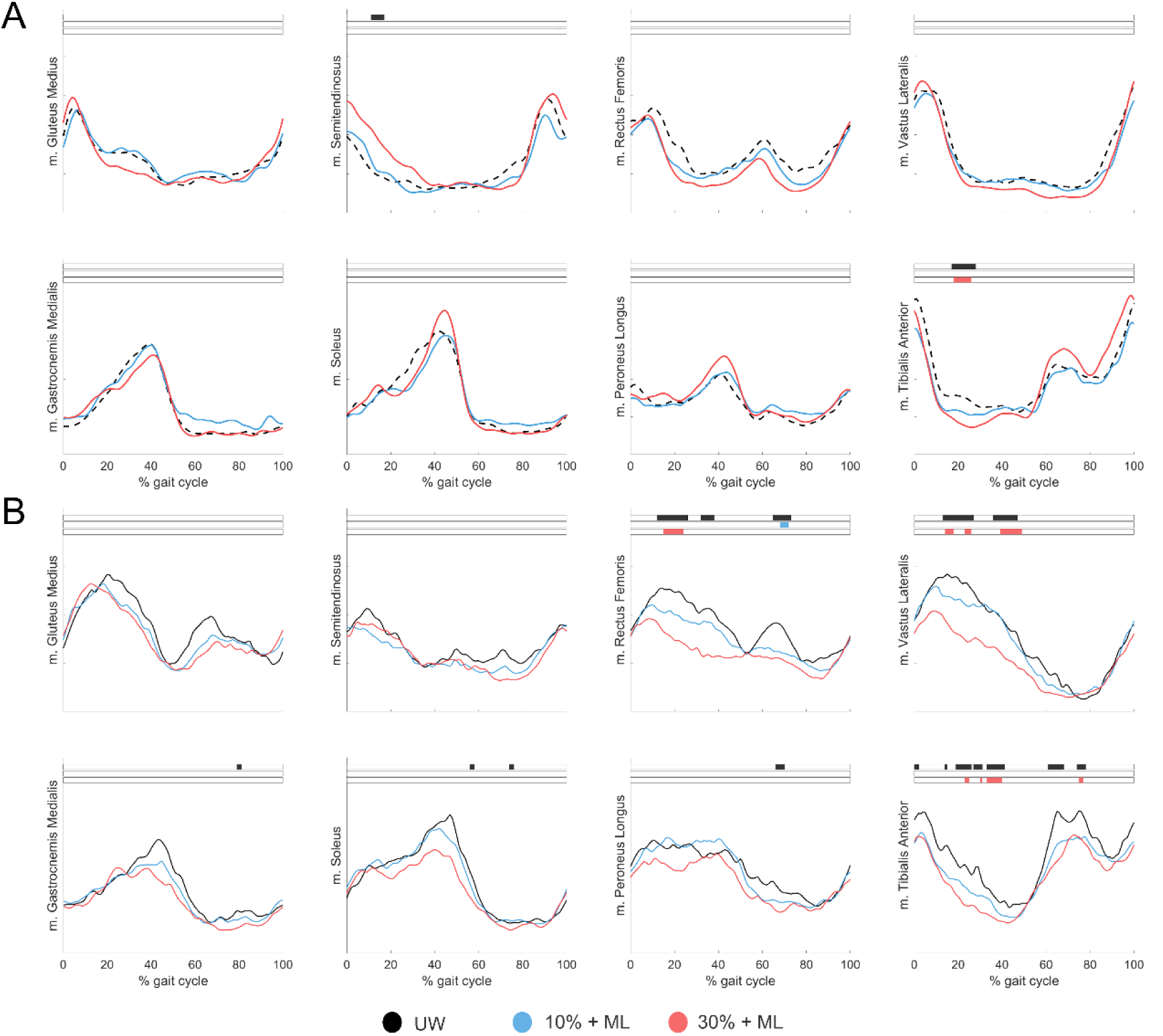
Mean muscle activation patterns across the gait cycle during walking with mediolateral (ML) forces for *A)* able-bodied individuals and *B)* individuals after stroke during Unsupported Walking (UW; black), 10% BWS + ML (blue) and 30% BWS + ML (red). Black (main effect) and colored (post-hoc effect) boxes show the part of the gait cycle in which a significant difference is present (p<0.05).

For individuals after stroke (Figure 4), decreased m. Rectus Femoris activity during initial swing with 10%+ML (p=0.01) and during midstance with 30%+ML (p<0.001) were found. For m. Vastus Lateralis, a decrease in activity with 30%+ML (p=0.01, p=0.01, p<0.001) was found. For m. Tibialis Anterior, decreased activity during stance (p=0.02, p=0.04, p<0.001) and initial swing (p=0.03) with 30%+ML were found.

### Body weight support with anteroposterior forces

In this section, conditions will be presented as BWS% ± AP%. There was an increase in walking speed with 10%+1AP, 10%+2AP, 30%+1AP, and 30%+2AP (all p<0.001, Figure 2). Significant condition*group interactions were found for all conditions (all p<0.001) except for 30%+1AP, showing that in three out of four conditions with anterior forces, there was an increase in speed in individuals after stroke, but a decrease or smaller increase in able-bodied individuals. In addition, in all conditions with posterior forces, there was a decrease in speed for able-bodied controls and an increase for individuals after stroke. Step length increased in all conditions, except for 10%-2AP (all p<0.05). A significant condition*least affected leg interaction was found for 30%+2AP, indicating that step length increased more for the least affected compared to the most affected leg. Significant condition*able-bodied leg interactions were found for 10%-1AP, 10%-2AP and 30%-1AP (all p<0.05), indicating that step length increased for the most affected leg after stroke, but decreased in able-bodied individuals. Single stance time increased with 30%+2AP compared to UW (p<0.001). For step width, no significant differences between groups or conditions were found. Step width variability was larger in able-bodied individuals than in individuals after stroke (p=0.02) and depended on age (p=0.04). Step width variability increased with 30%+2AP compared to UW (p=0.03).

For able-bodied individuals (Figure 5), decreased m. Gluteus Medius activity during terminal stance was found with 30%+1AP (p=0.003). For m. Semitendinosus, increased activity during early stance was found with 30%+1AP, 30%-1AP and 30%+2AP (all p <0.001). For m. Gastrocnemius Medialis, we found decreased activity with 10%-2AP (p=0.03) and increased activity at 30%+2AP (p=0.04), both during early stance. For m. Soleus, we found decreased activity with 10%-2AP (p<0.001) and increased activity with 30%+2AP (p=0.004), both during early stance. During late stance, activity was increased with 30%-2AP (p=0.03). For m. Peroneus Longus, increased activity during late stance was found with 30%-2AP (p=0.003).

**Figure 5.**
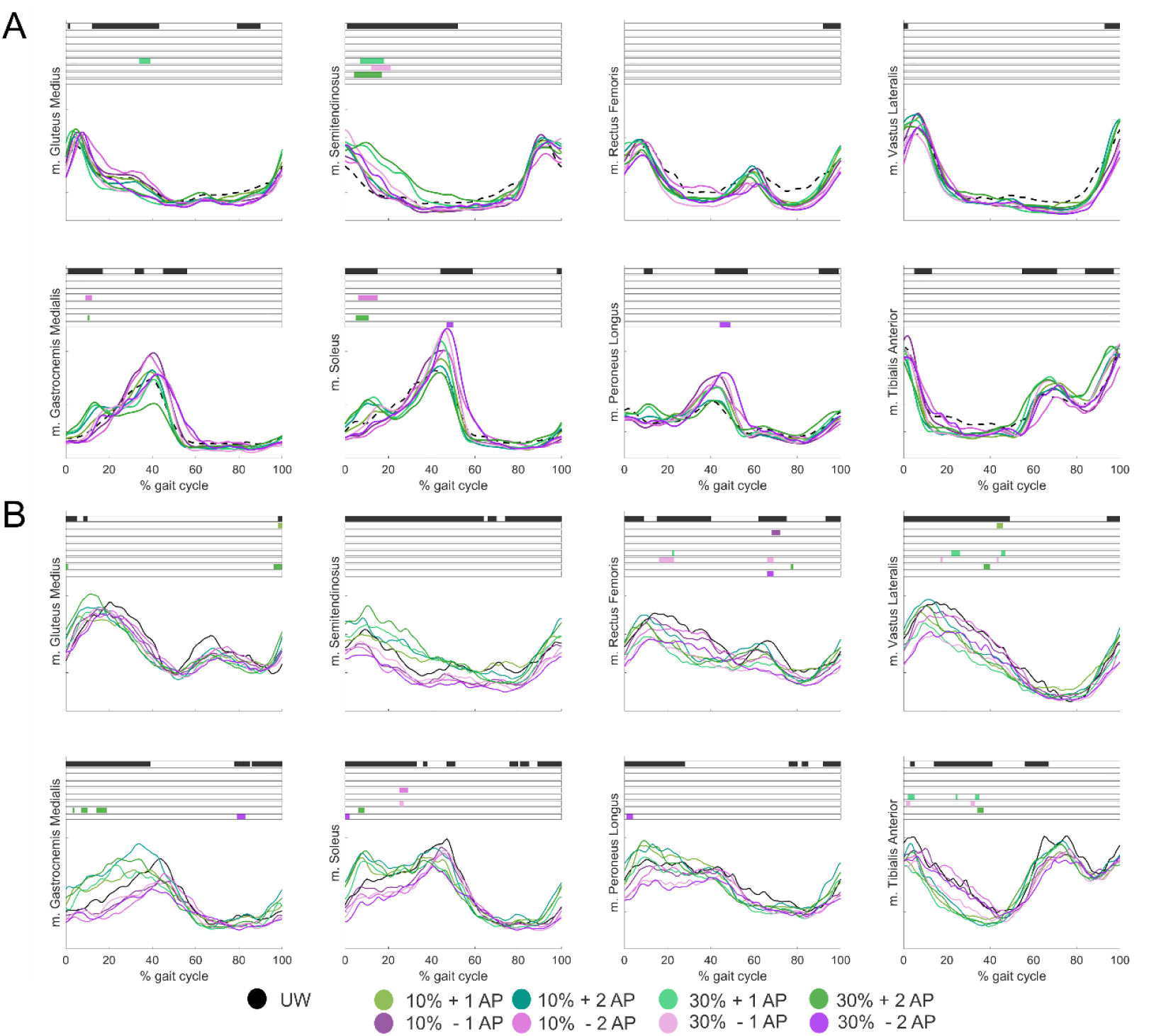
Mean muscle activation patterns across the gait cycle during walking with anteroposterior (AP) forces for *A)* able-bodied individuals and *B)* individuals after stroke during Unsupported Walking (UW; black), anterior forces (green) and posterior forces (purple). Black (main effect) and colored (post-hoc effect) boxes show the part of the gait cycle in which a significant difference is present (p<0.05).

For individuals after stroke (Figure 5), increased m. Gluteus Medius activity during terminal stance was found with 10%+1AP (p=0.03) and 30%+2AP (p=0.04, p=0.03, p=0.04). For m. Rectus Femoris, decreased activity was found with 10%-1AP (initial swing, p=0.01), 30%+1AP (midstance, p=0.04), 30%-1AP (midstance p=0.001, initial swing p=0.02), 30%+2AP (initial swing, p=0.04) and 30%-2AP (initial swing, p=0.02). For m. Vastus Lateralis, decreased activity was found with 10%+1AP (midstance, p=0.02), 30%+1AP (late stance, p=0.01, p=0.04), 30%-1AP (early and late stance, both p=0.04) and 30%+2AP (late stance, p=0.03). For m. Gastrocnemius, increased activity was found with 30%+2AP (loading response, p=0.03, p=0.01, p=0.001) and decreased activity with 30%-2AP (initial swing, p=0.01). For m. Soleus, decreased activity was found with 10%-2AP (midstance, p=0.002), 30%-1AP (midstance, p=0.03) and 30%+2AP (loading response, p=0.01) and increased activity was found with 30%-2AP (loading response, p=0.02). For m. Peroneus Longus, decreased activity was found with 30%-2% (loading response, p=0.02). For m. Tibialis Anterior, decreased activity was found with 30%+1AP (early stance, p=0.02, p=0.03, p=0.03), 30%-1AP (early stance, p=0.02, p=0.04, p=0.04, p=0.04), 30%+2AP (midstance, p=0.02, p=0.04) and 30%-2AP (early swing, p=0.04).

### Confidence in balance control

Individuals after stroke had increased confidence in balance control (Appendix C) compared to UW (74.0 ± 20.0) in the following conditions: 10% BWS (80.3 ± 17.5, p=0.02), 10%+ML (82.5 ± 15.4, p=0.01), 30%+ML (81.7 ± 16.3, p=0.03), 10%+1AP (80.7 ± 16.9, p=0.02), 10%-1AP (81.7 ± 18.0, p=0.02), 30%+1AP (80.7 ± 16.2, p=0.01) and 30%-1AP (81.7 ± 16.5, p=0.03).

## Discussion

This study aimed to identify the effects of RYSEN BWS with and without the addition of mediolateral and anteroposterior forces on step parameters and muscular control in individuals after stroke and able-bodied individuals. We formulated specific hypotheses for all RYSEN-induced forces, which will be discussed point-by-point in the sections below.

Our results show that BWS affects gait speed and stability differently in individuals after stroke and able-bodied individuals. Similar to previous research (35), walking speed increased in individuals after stroke but decreased in able-bodied individuals. Increased walking speeds in individuals after stroke may be attributed to the supportive properties of BWS devices (35) and to enhanced paretic propulsion and decreased non-paretic braking impulses (36). They may also result from enhanced confidence in balance control when walking in the RYSEN (37). A reasonable explanation for decreased walking speed in able-bodied individuals with BWS could be reduced voluntary effort when walking with BWS.

Contrary to our expectations, we found that BWS reduced step width variability and m. Gluteus Medius activity in able-bodied individuals, but not in individuals after stroke. Both reductions are associated with improved gait stability (14, 25, 38). It is known that, on the one hand, BWS can improve gait stability by providing additional sensory information on position as BWS acts as an external reference point (25). On the other hand, BWS can reduce gait stability by decreasing the gravitational moment about the stance limb (25), which normally helps to stabilize by balancing the whole body’s lateral inertial moment (16). Since different results were found for both groups, our findings may suggest that BWS benefits able-bodied individuals but impairs individuals after stroke, as their increased step width variability could indicate reduced stability. This may indicate that the BWS force could be considered a perturbation rather than support and that improved speed and balance confidence in individuals after stroke mainly result from the fall prevention mechanism. Our results may indicate that individuals after stroke, unlike able-bodied individuals, exhibit limited adaptive capacity to turn the BWS force into a supportive feature to enhance stability.

We hypothesized that adding mediolateral forces to BWS would further enhance frontal plane stability, but our results did not support this. Apart from a decrease in tibialis anterior activity in both groups, suggesting a lower need for active balance control via the ankle strategy (39), we did not find signs of improved stability in the BWS+ML conditions. This suggests that mediolateral forces of the RYSEN do not act as a stabilizing mechanism in the frontal plane as we expected. This could be because the mediolateral forces act on the overhead sling instead of the pelvis, as in previous studies (15, 40). As such, the device still allows for lateral movements and might not stabilize as effectively as pelvis stabilization (15), holding handrails (41) or other stabilizers. Previous work (27) on mediolateral forces combined with BWS also showed limited differences in gait stability indicators compared to BWS only. It needs further investigation to determine whether mediolateral forces of the RYSEN can be of added value during more challenging dynamic balance tasks or for individuals that show large lateral excursions during walking, for instance in ataxia or pusher syndrome.

Our hypotheses for anteroposterior forces were partly confirmed. We found the expected increase in walking speed with anterior forces, but not the expected increase in push-off capacity with posterior forces. All anteroposterior forces increased walking speed in individuals after stroke, but the increases were most pronounced with anterior forces. Similar to previous work (42), the increase in walking speed after stroke was accompanied by step length increases of the least affected leg. While this might suggest that push-off power of the most affected leg increased, it is more likely attributable to the assistance of the anterior forces, as plantar flexor activity of the most affected leg did not increase during push-off. Therefore, other strategies likely contributed to increased walking speed, for example, adopting a forward-leaning posture (10) to use gravity for propulsion, increasing hip flexor activity (43) or improving knee flexion during push-off due to enhanced stability (44). These strategies, instead of increasing push-off capacity as expected, could also have been used to counteract the posterior forces. While slightly increased m. Rectus Femoris activity during these conditions may support increased hip flexor activity, further research, including kinetic and kinematic assessments of gait, needs to determine how posterior forces can be better utilized during gait training to enhance propulsion capacity in individuals after stroke.

Besides the anticipated effects on muscular control as mentioned in our hypotheses, we observed several other changes that are worth mentioning. With BWS, BWS+ML and BWS+AP, decreased m. Rectus Femoris and m. Vastus Lateralis activity was found in individuals after stroke. Decreased knee extensor activity could result from decreased body weight and a smaller knee flexion angle at foot-flat with BWS (45). In addition, we found increased semitendinosus activity with anterior forces, mostly pronounced in individuals after stroke. This might be an eccentric contraction, generating backward momentum to brake the forward movement. Slightly increased plantar flexor activity during early stance with anterior forces may also reflect an eccentric contraction to brake the forward movement (46).

The findings of this study may have important consequences for clinical practice. First, we found increased step length, walking speed and balance confidence with BWS in individuals after stroke but differences compared to UW were otherwise limited. This suggests that the main effect of the RYSEN is to provide a safe and controlled gait training environment in which patients can practice walking at a relatively high speed with increased balance confidence. Many effects observed in the BWS+ML and BWS+AP conditions were equivalent to the effects in the BWS condition. Hence these effects seem to arise predominantly from vertical BWS. Adding mediolateral forces did not have the expected effect, as they had little effect on frontal plane stability in individuals after stroke. During normal walking conditions, as performed in this study, mediolateral forces rather seem to have a destabilizing effect and thus should not be recommended as facilitators of gait training when enhanced frontal plane stability is desired. Adding anterior forces can evoke increased walking speeds in individuals after stroke. In contrast, posterior forces did not have the expected effect on increasing propulsion capacity. When this effect is envisioned, potential extra instruction to the patient to utilize this feature could be required.

While this study provides important insights into the applicability of the RYSEN, there are some considerations. First, this study did not assess the isolated effect of mediolateral and anteroposterior forces, i.e. without BWS, since the RYSEN does not support this feature. Second, we need to acknowledge that the SPM analyses have an increased risk for type I errors because of the large number of comparisons. However, we reduced the chance of type I errors by only assessing post-hoc effects that overlapped with significant main effects. Third, it needs to be taken into account that individuals after stroke in this study were relatively good walkers, as they had to be able to walk at least ten meters without any support. Therefore, it would be of interest to see if comparable results can be found in a group of more impaired individuals after stroke, as they are considered a very relevant target group of the RYSEN. Finally, the data were analyzed at group level, which might have obscured individual differences in this heterogeneous stroke population, where some cases showed more pronounced responses than others. Future research may explore which clinical characteristics distinguish responders from non-responders.

## Conclusions

This study assessed to what extent different levels of BWS with and without mediolateral and anteroposterior forces of the RYSEN affected step parameters and muscular control in individuals after stroke and able-bodied individuals. Our results show that RYSEN BWS allows for safe gait training at a higher speed than unsupported walking and with increased balance confidence for patients. The added effects of mediolateral and anteroposterior forces in individuals after stroke were modest, currently offering limited evidence to support their use in targeted gait training. It should be explored how their potential can be better utilized during gait training, for instance with additional instructions, or different and more challenging gait tasks.

## Data Availability

All data produced in the present study are available upon reasonable request to the authors.

## Acknowledgements

The authors would like to thank the participants for their time and willingness to take part in this study. Furthermore, the authors thank Djarna Bijvoet, Geertje Pennink and Sophie Oberink for their contribution to the data collection, and Richard Fickert, Wim Kaan, Dirk van der Meer, and Emyl Smid for their technical support.

## Author Approval

All authors have seen and approved the manuscript.

### Appendix A

STROBE Statement—Checklist of items that should be included in reports of ***cross-sectional studies***

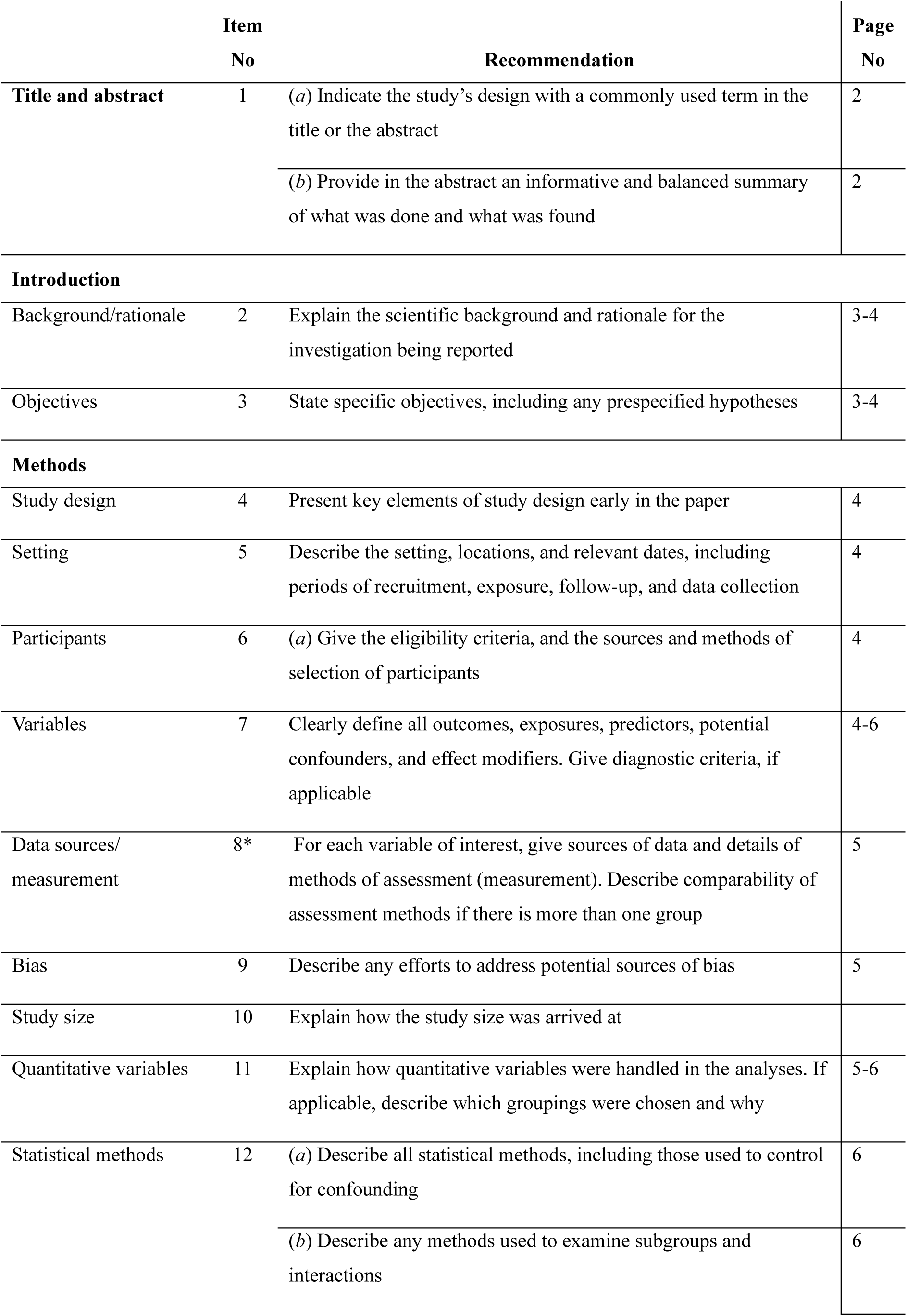
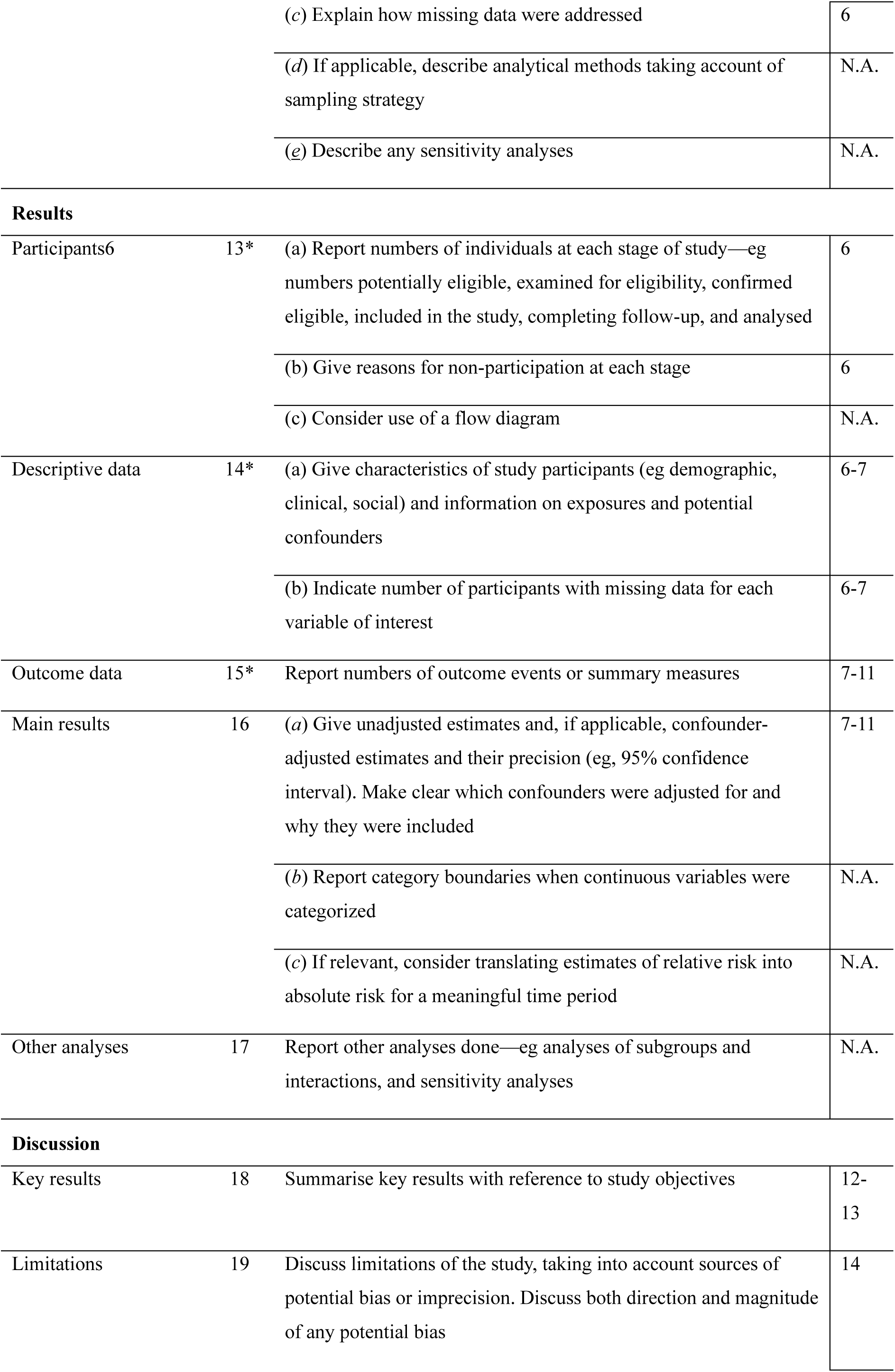
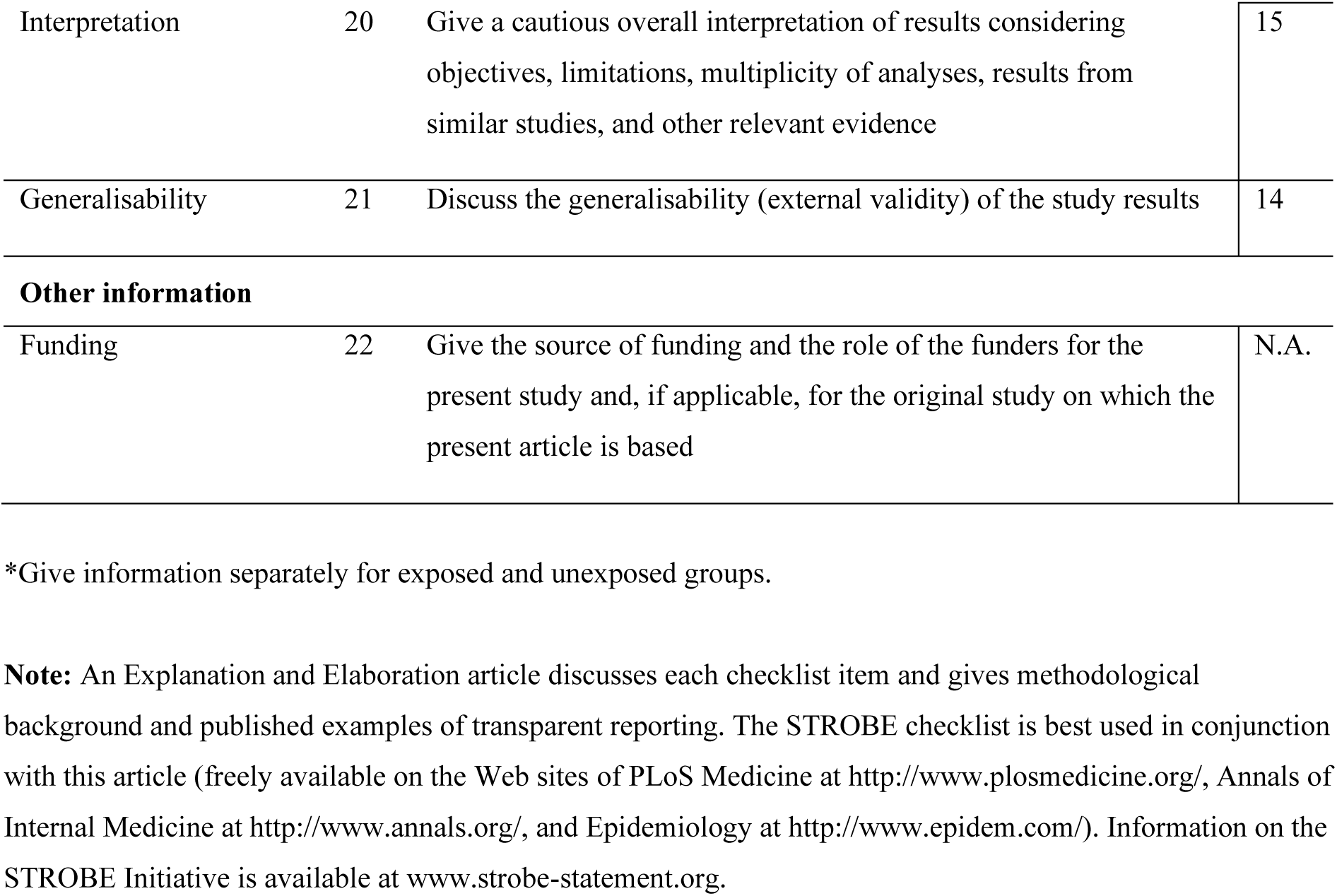

### Appendix B

| Body weight support (BWS) |  |  |  |  |  |
| --- | --- | --- | --- | --- | --- |
|  | Step length | Single stance time | Walking speed | Step width | Step width variability |
| Intercept | $\beta=0.50$<br>$F(1,89)=4.64$<br>**$p<0.001$** | $\beta=0.45$<br>$F(1,102)=4.02$<br>**$p<0.001$** | $\beta=0.68$<br>$F(1,83)=3.31$<br>**$P<0.001$** | $\beta=0.15$<br>$F(1,70)=5.76$<br>**$P<0.001$** | $\beta=0.02$<br>$F(1,69)=1.19$<br>$P=0.12$ |
| Least affected leg | $\beta=-0.04$<br>$F(1,89)=-0.90$<br>$P=0.37$ | $\beta=0.08$<br>$F(1,102)=1.67$<br>$P=0.09$ | | | |
| Able-bodied leg | $\beta=0.34$<br>$F(1,89)=5.88$<br>**$P<0.001$** | $\beta=-0.07$<br>$F(1,102)=-1.15$<br>$P=0.25$ | $\beta=0.93$<br>$F(1,83)=9.28$<br>**$P<0.001$** | $\beta=-0.02$<br>$F(1,70)=-2.01$<br>**$P=0.04$** | $\beta=0.01$<br>$F(1,69)=1.21$<br>$P=0.09$ |
| Age | $\beta=-0.002$<br>$F(1,89)=-0.99$<br>$P=0.32$ | $\beta=0.001$<br>$F(1,102)=0.38$<br>$P=0.71$ | $\beta=-0.005$<br>$F(1,83)=-1.20$<br>$P=0.23$ | $\beta=-0.0002$<br>$F(1,70)=-0.49$<br>$P=0.62$ | $\beta=0.0002$<br>$F(1,69)=0.97$<br>$P=0.44$ |
| 10% BWS | $\beta=0.05$<br>$F(1,89)=3.27$<br>**$P<0.001$** | $\beta=-0.01$<br>$F(1,102)=-0.56$<br>$P=0.58$ | $\beta=0.07$<br>$F(1,83)=2.83$<br>**$P=0.01$** | $\beta=0.0011$<br>$F(1,70)=0.18$<br>$P=0.86$ | $\beta=0.005$<br>$F(1,69)=0.57$<br>$P=0.41$ |
| 30% BWS | $\beta=0.05$<br>$F(1,89)=3.15$<br>**$P<0.001$** | $\beta=0.07$<br>$F(1,102)=2.79$<br>**$P=0.01$** | $\beta=0.05$<br>$F(1,83)=2.16$<br>**$P=0.03$** | $\beta=0.0011$<br>$F(1,70)=0.19$<br>$P=0.85$ | $\beta=0.006$<br>$F(1,69)=1.64$<br>$P=0.30$ |
| Least affected leg* 10% BWS | $\beta=-0.02$<br>$F(1,89)=-0.74$<br>$P=0.46$ | $\beta=0.01$<br>$F(1,102)=0.33$<br>$P=0.74$ | | | |
| Able-bodied leg * 10% BWS | $\beta=-0.05$<br>$F(1,89)=-1.92$<br>$P=0.06$ | $\beta=0.04$<br>$F(1,102)=0.85$<br>$P=0.40$ | $\beta=-0.13$<br>$F(1,83)=-3.88$<br>**$P<0.001$** | $\beta=-0.007$<br>$F(1,70)=-0.66$<br>$P=0.51$ | $\beta=-0.03$<br>$F(1,69)=-1.27$<br>**$P=0.003$** |
| Least affected leg * 30% BWS | $\beta=0.03$<br>$F(1,89)=1.29$ | $\beta=-0.05$<br>$F(1,102)=-1.32$ | | | |
|  | P=0.20 | P=0.19 |  |  |  |
| Able-bodied leg * 30% BWS | $\beta=-0.03$<br>F(1,89)=-0.89<br>P=0.38 | $\beta=-0.05$<br>F(1,102)=-1.18<br>P=0.24 | $\beta=-0.04$<br>F(1,83)=-1.31<br>P=0.20 | $\beta=0.004$<br>F(1,70)=0.33<br>P=0.74 | $\beta=-0.02$<br>F(1,69)=-2.64<br>**P=0.02** |

| Mediolateral forces (ML) |  |  |  |  |  |
| --- | --- | --- | --- | --- | --- |
|  | Step length | Single stance time | Walking speed | Step width | Step width variability |
| Intercept | $\beta=0.53$<br>F(1,97)=5.00<br>**p<0.001** | $\beta=0.37$<br>F(1,105)=3.11<br>**p<0.001** | $\beta=0.70$<br>F(1,82)=3.29<br>**P<0.001** | $\beta=0.11$<br>F(1,69)=4.56<br>**P<0.001** | $\beta=0.03$<br>F(1,69)=2.20<br>**P=0.03** |
| Least affected leg | $\beta=-0.04$<br>F(1,97)=-0.88<br>P=0.38 | $\beta=0.08$<br>F(1,105)=1.59<br>P=0.11 | | | |
| Able-bodied leg | $\beta=0.35$<br>F(1,97)=6.07<br>**P<0.001** | $\beta=-0.08$<br>F(1,105)=-1.31<br>P=0.19 | $\beta=0.94$<br>F(1,82)=9.05<br>**P<0.001** | $\beta=-0.04$<br>F(1,69)=-2.72<br>**P=0.01** | $\beta=0.01$<br>F(1,69)=1.90<br>P=0.06 |
| Age | $\beta=-0.003$<br>F(1,97)=-1.32<br>P=0.19 | $\beta=0.002$<br>F(1,105)=1.02<br>P=0.31 | $\beta=-0.005$<br>F(1,82)=-1.23<br>P=0.22 | $\beta=-0.001$<br>F(1,69)=0.98<br>P=0.33 | B<0.001<br>F(1,69)=0.10<br>P=0.92 |
| 10% BWS | $\beta=0.011$<br>F(1,97)=0.68<br>P=0.50 | $\beta=0.07$<br>F(1,105)=2.06<br>**P=0.04** | $\beta=0.05$<br>F(1,82)=1.70<br>P=0.09 | $\beta=-0.01$<br>F(1,69)=-1.08<br>P=0.28 | $\beta=0.001$<br>F(1,69)=0.27<br>P=0.79 |
| 30% BWS | $\beta=0.04$<br>F(1,97)=2.42<br>**P=0.02** | $\beta=0.07$<br>F(1,105)=2.19<br>**P=0.03** | $\beta=0.07$<br>F(1,82)=2.54<br>**P=0.01** | $\beta=-0.01$<br>F(1,69)=-1.28<br>P=0.21 | $\beta=0.001$<br>F(1,69)=0.20<br>P=0.84 |
| Least affected leg * 10% BWS + ML | $\beta=0.02$<br>F(1,97)=1.00<br>P=0.32 | $\beta=-0.07$<br>F(1,105)=-1.70<br>P=0.09 | | | |
| Able-bodied leg * 10%<br>BWS + ML | $\beta=-0.03$<br>$F(1,97)=-0.95$<br>$P=0.34$ | $\beta=-0.05$<br>$F(1,105)=-0.94$<br>$P=0.35$ | $\beta=-0.10$<br>$F(1,82)=-2.39$<br>**P=0.02** | $\beta=0.002$<br>$F(1,69)=0.15$<br>$P=0.88$ | $\beta=-0.02$<br>$F(1,69)=-1.75$<br>$P=0.08$ |
| Least affected leg * 30%<br>BWS + ML | $\beta=0.02$<br>$F(1,97)=0.89$<br>$P=0.37$ | $\beta=-0.07$<br>$F(1,105)=-1.53$<br>$P=0.13$ | | | |
| Able-bodied leg * 30%<br>BWS + ML | $\beta=-0.04$<br>$F(1,97)=-1.43$<br>$P=0.16$ | $\beta=-0.04$<br>$F(1,105)=-0.70$<br>$P=0.48$ | $\beta=-0.11$<br>$F(1,82)=-2.74$<br>**P=0.01** | $\beta=0.02$<br>$F(1,69)=1.71$<br>$P=0.09$ | $\beta=-0.01$<br>$F(1,69)=-0.81$<br>$P=0.42$ |

| Anteroposterior forces (AP) |  |  |  |  |  |
| --- | --- | --- | --- | --- | --- |
|  | Step length | Single stance time | Walking speed | Step width | Step width variability |
| Intercept | $\beta=0.48$<br>$F(1,264)=3.84$<br>**p<0.001** | $\beta=0.43$<br>$F(1,293)=4.88$<br>**p<0.001** | $\beta=0.66$<br>$F(1, 251)=3.36$<br>**P<0.001** | $\beta=0.17$<br>$F(1,202)=5.53$<br>**P<0.001** | $\beta=0.06$<br>$F(1,202)=3.57$<br>**P<0.001** |
| Least affected leg | $\beta=-0.02$<br>$F(1,264)=-0.66$<br>$P=0.51$ | $\beta=0.08$<br>$F(1,293)=1.80$<br>$P=0.07$ | | | |
| Able-bodied leg | $\beta=0.32$<br>$F(1,264)=5.05$<br>**P<0.001** | $\beta=-0.06$<br>$F(1,293)=-1.20$<br>$P=0.23$ | $\beta=0.93$<br>$F(1, 251)=9.45$<br>**P<0.001** | $\beta=-0.02$<br>$F(1, 202)=-1.14$<br>$P=0.25$ | $\beta=0.03$<br>$F(1,202)=2.37$<br>**P=0.02** |
| Age | $\beta=-0.002$<br>$F(1,264)=-0.68$<br>$P=0.49$ | $\beta=0.001$<br>$F(1,293)=0.68$<br>$P=0.50$ | $\beta=-0.004$<br>$F(1, 251)=-1.13$<br>$P=0.26$ | $\beta=-0.001$<br>$F(1, 202)=-1.07$<br>$P=0.28$ | $\beta=-0.001$<br>$F(1,202)=-2.08$<br>**P=0.04** |
| 10% BWS + 1% AP | $\beta=0.05$<br>$F(1,264)=3.06$<br>**P=0.002** | $\beta=0.03$<br>$F(1,293)=0.98$<br>$P=0.33$ | $\beta=0.13$<br>$F(1, 251)=3.87$<br>**P<0.001** | $\beta=0.002$<br>$F(1, 202)=0.24$<br>$P=0.81$ | $\beta=0.01$<br>$F(1,202)=1.15$<br>$P=0.25$ |
| 10% BWS -1% AP | $\beta=0.04$<br>$F(1,264)=2.25$ | $\beta=0.04$<br>$F(1,293)=1.34$ | $\beta=0.03$<br>$F(1, 251)=0.94$ | $\beta=0.004$<br>$F(1, 202)=0.46$ | $\beta=0.01$<br>$F(1,202)=0.73$ |
|  | **P=0.03** | P=0.18 | P=0.35 | P=0.65 | P=0.47 |
| 10% BWS +2% AP | $\beta=0.06$<br>F(1,264)=3.47<br>**P<0.001** | $\beta=0.01$<br>F(1,293)=0.30<br>P=0.76 | $\beta=0.11$<br>F(1, 251)=3.34<br>**P<0.001** | $\beta=-0.001$<br>F(1, 202)=-0.13<br>P=0.90 | $\beta=0.01$<br>F(1,202)=1.46<br>P=0.14 |
| 10% BWS -2% AP | $\beta=0.03$<br>F(1,264)=1.86<br>P=0.06 | $\beta=0.05$<br>F(1,293)=1.60<br>P=0.11 | $\beta=0.002$<br>F(1, 251)=0.05<br>P=0.96 | $\beta=0.001$<br>F(1, 202)=0.15<br>P=0.88 | $\beta=0.001$<br>F(1,202)=0.16<br>P=0.88 |
| 30% BWS +1% AP | $\beta=0.06$<br>F(1,264)=3.40<br>**P<0.001** | $\beta=0.02$<br>F(1,293)=0.53<br>P=0.60 | $\beta=0.13$<br>F(1, 251)=3.93<br>**P<0.001** | $\beta=0.003$<br>F(1, 202)=0.38<br>P=0.70 | $\beta=0.01$<br>F(1,202)=1.10<br>P=0.27 |
| 30% BWS -1% AP | $\beta=0.07$<br>F(1,264)=4.06<br>**P<0.001** | $\beta=0.04$<br>F(1,293)=1.25<br>P=0.21 | $\beta=0.06$<br>F(1, 251)=1.80<br>P=0.07 | $\beta=0.005$<br>F(1, 202)=0.61<br>P=0.54 | $\beta=0.01$<br>F(1,202)=0.73<br>P=0.47 |
| 30% BWS +2% AP | $\beta=0.05$<br>F(1,264)=3.21<br>**P=0.001** | $\beta=0.03$<br>F(1,293)=1.06<br>P=0.29 | $\beta=0.14$<br>F(1, 251)=4.35<br>**P<0.001** | $\beta=0.007$<br>F(1, 202)=0.93<br>P=0.35 | $\beta=0.02$<br>F(1,202)=2.16<br>**P=0.03** |
| 30% BWS -2% AP | $\beta=0.03$<br>F(1,264)=2.01<br>**P=0.04** | $\beta=0.11$<br>F(1,293)=3.84<br>**P<0.001** | $\beta=0.01$<br>F(1, 251)=0.35<br>P=0.72 | $\beta=0.005$<br>F(1, 202)=0.61<br>P=0.54 | $\beta=0.01$<br>F(1,202)=0.80<br>P=0.42 |
| Least affected leg * 10%<br>BWS + 1% AP | $\beta=0.03$<br>F(1,264)=1.18<br>P=0.24 | $\beta=-0.05$<br>F(1,293)=-1.06<br>P=0.28 | | | |
| Able-bodied leg * 10%<br>BWS + 1% AP | $\beta=-0.02$<br>F(1,264)=-0.77<br>P=0.44 | $\beta=-0.01$<br>F(1,293)=-0.29<br>P=0.77 | $\beta=-0.12$<br>F(1, 251)=-2.62<br>**P=0.009** | $\beta=0.01$<br>F(1, 202)=0.73<br>P=0.47 | $\beta=-0.003$<br>F(1,202)=-0.27<br>P=0.79 |
| Least affected leg * 10%<br>BWS -1% AP | $\beta=-0.004$<br>F(1,264)=-0.16<br>P=0.87 | $\beta=-0.02$<br>F(1,293)=-0.46<br>P=0.65 | | | |
| Able-bodied leg * 10%<br>BWS -1% AP | $\beta=-0.07$<br>F(1,264)=-2.21<br>**P=0.03** | $\beta=-0.03$<br>F(1,293)=-0.58<br>P=0.56 | $\beta=-0.14$<br>F(1, 251)=-3.12<br>**P=0.002** | $\beta=-0.01$<br>F(1, 202)=-0.51<br>P=0.61 | $\beta=-0.02$<br>F(1,202)=-1.36<br>P=0.17 |
| Least affected leg * 10%<br>BWS + 2% AP | $\beta=0.03$<br>F(1,264)=1.07<br>P=0.29 | $\beta=-0.03$<br>F(1,293)=-0.68<br>P=0.50 | | | |
| Able-bodied leg * 10%<br>BWS + 2% AP | $\beta=-0.04$<br>F(1,264)=-0.18<br>P=0.24 | $\beta<-0.0001$<br>F(1,293)=-0.0004<br>P=0.99 | $\beta=-0.14$<br>F(1, 251)=-2.95<br>**P=0.003** | $\beta=0.003$<br>F(1, 202)=0.24<br>P=0.81 | $\beta=-0.02$<br>F(1,202)=-1.50<br>P=0.13 |
| Least affected leg * 10%<br>BWS - 2% AP | $\beta=-0.08$<br>F(1,264)=-1.58<br>P=0.12 | $\beta=-0.07$<br>F(1,293)=-1.56<br>P=0.12 | | | |
| Able-bodied leg * 10%<br>BWS -2% AP | $\beta=0.008$<br>F(1,264)=-2.30<br>**P=0.02** | $\beta=0.0003$<br>F(1,293)=0.006<br>P=0.99 | $\beta=-0.29$<br>F(1, 251)=-6.33<br>**P<0.001** | $\beta=0.004$<br>F(1, 202)=0.31<br>P=0.76 | $\beta=0.0002$<br>F(1,202)=0.02<br>P=0.98 |
| Least affected leg * 30%<br>BWS + 1% AP | $\beta=-0.04$<br>F(1,264)=0.28<br>P=0.78 | $\beta=-0.03$<br>F(1,293)=-0.68<br>P=0.50 | | | |
| Able-bodied leg * 30%<br>BWS + 1% AP | $\beta=-0.04$<br>F(1,264)=-1.18<br>P=0.24 | $\beta=0.001$<br>F(1,293)=0.02<br>P=0.98 | $\beta=-0.08$<br>F(1, 251)=-1.74<br>P=0.08 | $\beta=0.01$<br>F(1, 202)=0.64<br>P=0.52 | $\beta=-0.02$<br>F(1,202)=-1.60<br>P=0.11 |
| Least affected leg * 30%<br>BWS - 1% AP | $\beta=-0.01$<br>F(1,264)=-0.57<br>P=0.57 | $\beta=-0.01$<br>F(1,293)=-0.24<br>P=0.81 | | | |
| Able-bodied leg * 30%<br>BWS -1% AP | $\beta=-0.08$<br>F(1,264)=-2.55<br>**P=0.01** | $\beta=-0.005$<br>F(1,293)=-0.10<br>P=0.92 | $\beta=-0.14$<br>F(1, 251)=-2.97<br>**P=0.003** | $\beta=0.02$<br>F(1, 202)=1.43<br>P=0.16 | $\beta=-0.01$<br>F(1,202)=-0.71<br>P=0.48 |
| Least affected leg * 30%<br>BWS + 2% AP | $\beta=0.06$<br>F(1,264)=2.43<br>**P=0.02** | $\beta=-0.08$<br>F(1,293)=-1.83<br>P=0.07 | | | |
| Able-bodied leg * 30%<br>BWS + 2% AP | $\beta=0.003$<br>F(1,264)=0.11<br>P=0.91 | $\beta=-0.003$<br>F(1,293)=-0.08<br>P=0.94 | $\beta=-0.10$<br>F(1, 251)=-2.11<br>**P=0.04** | $\beta=0.01$<br>F(1, 202)=1.04<br>P=0.30 | $\beta=0.01$<br>F(1,202)=1.27<br>P=0.21 |
| Least affected leg * 30%<br>BWS - 2% AP | $\beta=-0.01$<br>F(1,264)=-0.54 | $\beta=-0.06$<br>F(1,293)=-1.37 | | | |
|  | P=0.59 | P=0.17 |  |  |  |
| Able-bodied leg *30%<br>BWS – 2% AP | $\beta=-0.04$<br>F(1,264)=-1.19<br>P=0.23 | $\beta=-0.06$<br>F(1,293)=-1.35<br>P=0.17 | $\beta=-0.15$<br>F(1, 251)=-3.31<br>**P=0.001** | $\beta=0.01$<br>F(1, 202)=1.04<br>P=0.30 | $\beta=-0.02$<br>F(1,202)=-1.41<br>P=0.16 |

### Appendix C. Questionnaire on confidence in maintaining balance and avoiding a fall (31)

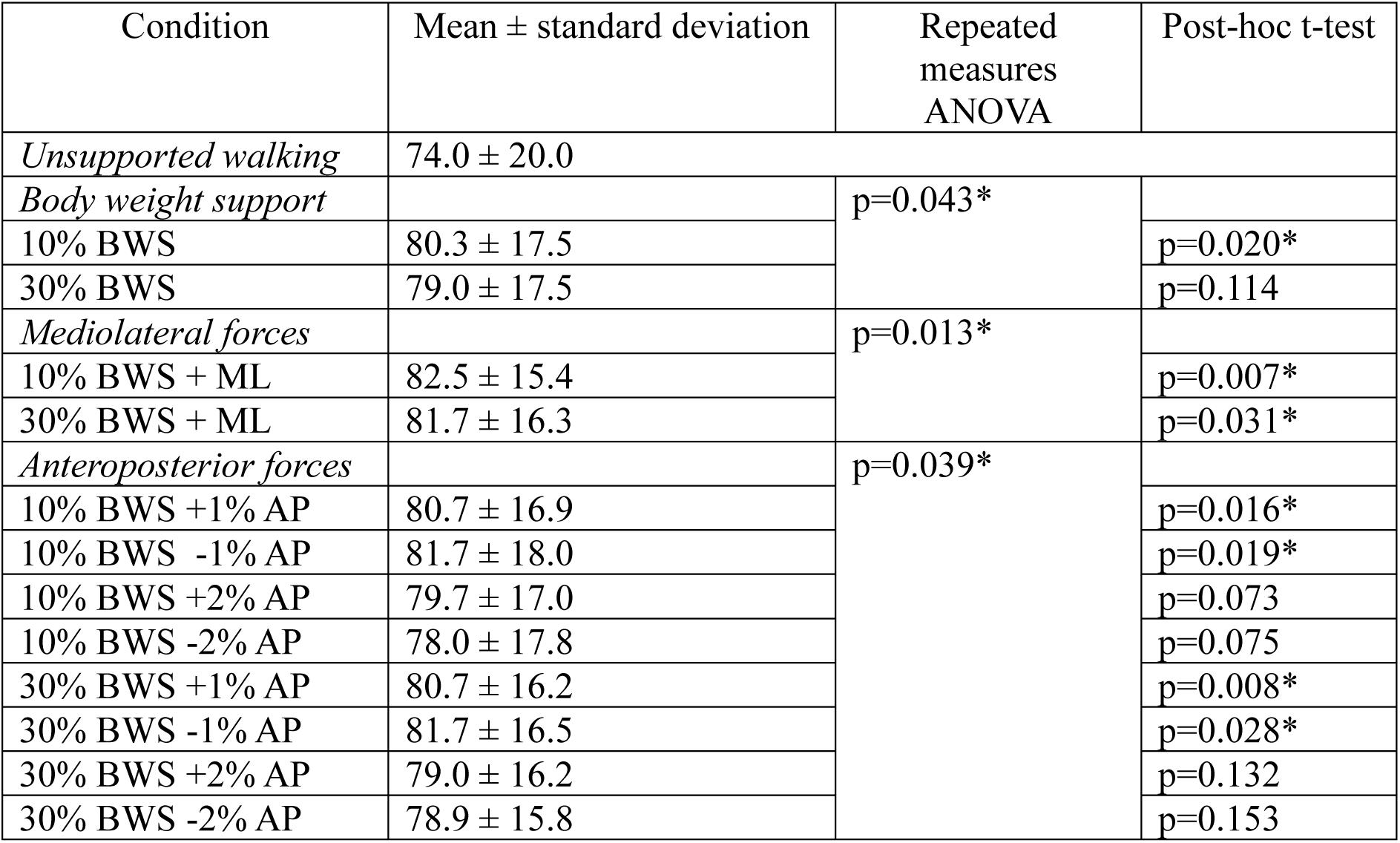

## References

1. Mehrholz J, Thomas S, Elsner B. Treadmill training and body weight support for walking after stroke. Cochrane Database of Systematic Reviews. 2017;2017(8).

2. Ettema S, Pennink GH, Buurke TJ, David S, van Bennekom CA, Houdijk H. Clinical indications and protocol considerations for selecting initial body weight support levels in gait rehabilitation: a systematic review. Journal of NeuroEngineering and Rehabilitation. 2024;21(1):97.

3. Plooij M, Keller U, Sterke B, Komi S, Vallery H, Von Zitzewitz J. Design of RYSEN: an intrinsically safe and low-power three-dimensional Overground body weight support. IEEE Robotics and Automation Letters. 2018;3(3):2253–60.

4. Barela AMF, Gama GL, Russo DV, Celestino ML, Barela JA. Gait alterations during walking with partial body weight supported on a treadmill and over the ground. SCIENTIFIC REPORTS. 2019;9.

5. Fenuta AM, Hicks AL. Muscle activation during body weight-supported locomotion while using the ZeroG. Journal of Rehabilitation Research and Development. 2014;51(1):51–8.

6. Awai L, Franz M, Easthope C, Vallery H, Curt A, Bolliger M. Preserved gait kinematics during controlled body unloading. Journal of neuroengineering and rehabilitation. 2017;14(1):1–10.

7. Sousa CO, Barela JA, Prado-Medeiros CL, Salvini TF, Barela AM. The use of body weight support on ground level: an alternative strategy for gait training of individuals with stroke. Journal of neuroengineering and rehabilitation. 2009;6(1):1–10.

8. Apte S, Plooij M, Vallery H. Simulation of human gait with body weight support: benchmarking models and unloading strategies. Journal of neuroengineering and rehabilitation. 2020;17(1):81.

9. Apte S, Plooij M, Vallery H. Influence of body weight unloading on human gait characteristics: A systematic review. Journal of NeuroEngineering and Rehabilitation. 2018;15(1).

10. Plooij M, Apte S, Keller U, Baines P, Sterke B, Asboth L, et al. Neglected physical human-robot interaction may explain variable outcomes in gait neurorehabilitation research. SCIENCE ROBOTICS. 2021;6(58).

11. Donelan JM, Shipman DW, Kram R, Kuo AD. Mechanical and metabolic requirements for active lateral stabilization in human walking. Journal of biomechanics. 2004;37(6):827–35.

12. Bauby CE, Kuo AD. Active control of lateral balance in human walking. Journal of biomechanics. 2000;33(11):1433–40.

13. Kuo AD, Donelan JM. Dynamic principles of gait and their clinical implications. Physical therapy. 2010;90(2):157–74.

14. Dean JC, Alexander NB, Kuo AD. The effect of lateral stabilization on walking in young and old adults. IEEE Transactions on Biomedical Engineering. 2007;54(11):1919–26.

15. Ijmker T, Houdijk H, Lamoth CJ, Beek PJ, van der Woude LH. Energy cost of balance control during walking decreases with external stabilizer stiffness independent of walking speed. Journal of biomechanics. 2013;46(13):2109–14.

16. MacKinnon CD, Winter DA. Control of whole body balance in the frontal plane during human walking. Journal of biomechanics. 1993;26(6):633–44.

17. Maxwell Donelan J, Kram R, Arthur D K. Mechanical and metabolic determinants of the preferred step width in human walking. Proceedings of the Royal Society of London Series B: Biological Sciences. 2001;268(1480):1985-92.

18. Gottschall JS, Kram R. Energy cost and muscular activity required for propulsion during walking. Journal of Applied Physiology. 2003;94(5):1766–72.

19. Penke K, Scott K, Sinskey Y, Lewek MD. Propulsive forces applied to the body’s center of mass affect metabolic energetics poststroke. Archives of physical medicine and rehabilitation. 2019;100(6):1068–75.

20. Ellis RG, Sumner BJ, Kram R. Muscle contributions to propulsion and braking during walking and running: insight from external force perturbations. Gait & posture. 2014;40(4):594–9.

21. Dewolf A, Ivanenko YP, Mesquita RM, Lacquaniti F, Willems P. Neuromechanical adjustments when walking with an aiding or hindering horizontal force. European journal of applied physiology. 2020;120:91–106.

22. Weerdesteijn V, Niet Md, Van Duijnhoven H, Geurts AC. Falls in individuals with stroke. J Rehabil Res Dev. 2009;45(8):1195–214.

23. Von Schroeder HP, Coutts RD, Lyden PD, Billings E, Nickel VL. Gait parameters following stroke: a practical assessment. Journal of rehabilitation research and development. 1995;32:25-.

24. Turns LJ, Neptune RR, Kautz SA. Relationships between muscle activity and anteroposterior ground reaction forces in hemiparetic walking. Archives of physical medicine and rehabilitation. 2007;88(9):1127–35.

25. Dragunas AC, Gordon KE. Body weight support impacts lateral stability during treadmill walking. JOURNAL OF BIOMECHANICS. 2016;49(13):2662–8.

26. IJmker T, Lamoth C, Houdijk H, Tolsma M, Van Der Woude L, Daffertshofer A, et al. Effects of handrail hold and light touch on energetics, step parameters, and neuromuscular activity during walking after stroke. Journal of neuroengineering and rehabilitation. 2015;12:1–12.

27. Bannwart M, Bayer SL, König Ignasiak N, Bolliger M, Rauter G, Easthope CA. Mediolateral damping of an overhead body weight support system assists stability during treadmill walking. J Neuroeng Rehabil. 2020;17(1):108.

28. Faul F, Erdfelder E, Lang A-G, Buchner A. G* Power 3: A flexible statistical power analysis program for the social, behavioral, and biomedical sciences. Behavior research methods. 2007;39(2):175–91.

29. Association WM. World Medical Association Declaration of Helsinki: ethical principles for medical research involving human subjects. Jama. 2013;310(20):2191–4.

30. Hermens HJ, Freriks B, Merletti R, Stegeman D, Blok J, Rau G, et al. European recommendations for surface electromyography. Roessingh research and development. 1999;8(2):13–54.

31. Ellmers TJ, Cocks AJ, Young WR. Evidence of a link between fall-related anxiety and high-risk patterns of visual search in older adults during adaptive locomotion. The Journals of Gerontology: Series A. 2020;75(5):961–7.

32. Zeni Jr J, Richards J, Higginson J. Two simple methods for determining gait events during treadmill and overground walking using kinematic data. Gait & posture. 2008;27(4):710–4.

33. Õunpuu S, Winter DA. Bilateral electromyographical analysis of the lower limbs during walking in normal adults. Electroencephalography and clinical neurophysiology. 1989;72(5):429–38.

34. Pataky TC, Robinson MA, Vanrenterghem J. Region-of-interest analyses of one-dimensional biomechanical trajectories: bridging 0D and 1D theory, augmenting statistical power. PeerJ. 2016;4:e2652.

35. Burgess JK, Weibel GC, Brown DA. Overground walking speed changes when subjected to body weight support conditions for nonimpaired and post stroke individuals. Journal of neuroengineering and rehabilitation. 2010;7:6.

36. Hurt CP, Burgess JK, Brown DA. Limb contribution to increased self-selected walking speeds during body weight support in individuals poststroke. Gait and Posture. 2015;41(3):857–9.

37. Rosén E, Sunnerhagen KS, Kreuter M. Fear of falling, balance, and gait velocity in patients with stroke. Physiotherapy theory and practice. 2005;21(2):113–20.

38. van Hedel HJ, Rosselli I, Baumgartner-Ricklin S. Clinical utility of the over-ground bodyweight-supporting walking system Andago in children and youths with gait impairments. Journal of neuroengineering and rehabilitation. 2021;18:1–20.

39. Hof A, Duysens J. Responses of human ankle muscles to mediolateral balance perturbations during walking. Human movement science. 2018;57:69–82.

40. IJmker T, Houdijk H, Lamoth CJ, Jarbandhan AV, Rijntjes D, Beek PJ, et al. Effect of balance support on the energy cost of walking after stroke. Archives of physical medicine and rehabilitation. 2013;94(11):2255–61.

41. Buurke TJ, Lamoth CJ, van der Woude LH, den Otter R. Handrail holding during treadmill walking reduces locomotor learning in able-bodied persons. IEEE Transactions on Neural Systems and Rehabilitation Engineering. 2019;27(9):1753–9.

42. Ettema S, Blokland IJ, Buurke TJ, Houdijk H. The relationship between walking speed, step parameters, and margins of stability in individuals after stroke. Clinical Biomechanics. 2024.

43. Nadeau S, Gravel D, Arsenault AB, Bourbonnais D. Plantarflexor weakness as a limiting factor of gait speed in stroke subjects and the compensating role of hip flexors. Clinical biomechanics. 1999;14(2):125–35.

44. Kirtley C, Whittle MW, Jefferson R. Influence of walking speed on gait parameters. Journal of biomedical engineering. 1985;7(4):282–8.

45. Finch L, Barbeau H, Arsenault B. Influence of body weight support on normal human gait: development of a gait retraining strategy. Physical therapy. 1991;71(11):842–55.

46. Neptune RR, Kautz SA, Zajac FE. Contributions of the individual ankle plantar flexors to support, forward progression and swing initiation during walking. Journal of biomechanics. 2001;34(11):1387–98.

